# Can Koreans be ‘FREE’ from mask wearing?: Advanced mathematical model can suggest the idea

**DOI:** 10.1101/2023.01.03.23284126

**Authors:** Youngsuk Ko, Victoria May Mendoza, Renier Mendoza, Yubin Seo, Jacob Lee, Eunok Jung

**Affiliations:** Department of Mathematics, Konkuk University, Seoul, Korea; Institute of Mathematics, University of the Philippines Diliman, Quezon City, Philippines; Division of Infectious Disease, Department of Internal Medicine, Kangnam Sacred Heart Hospital, Hallym University College of Medicine, Seoul, Korea

## Abstract

**Background:** It was found that more than half of the population in Korea had a prior COVID-19 infection. In 2022, most nonpharmaceutical interventions, except mask-wearing indoors, had been lifted. Discussions about easing the indoor mask mandate are ongoing.

**Methods:** We developed an age-structured compartmental model that distinguishes vaccination history, prior infection, and medical staff from the rest of the population. Contact patterns among hosts were separated based on age and location. We simulated scenarios with the lifting of the mask mandate all at once or sequentially according to the locations. Furthermore, we investigated the impact of a new variant assuming that it has higher transmissibility and risk of breakthrough infection.

**Findings:** We found that the peak size of administered severe patients might not exceed 1,100 when the mask mandate is lifted everywhere, and 800 if the mask mandate only remains in the hospital. If the mask mandate is lifted in a sequence (except hospital), then the peak size of administered severe patients did not exceed 650. Moreover, if the new variant have both of higher transmissibility and immune reduction therefore the effective reproductive number of the new variant is approximately 3 times higher than the current variant, additional interventions may be needed to keep the administered severe patients from exceeding 2,000, which is the critical level we set.

**Interpretation:** Our findings showed that the lifting of the mask mandate, except in hospitals, would be applicable more manageable if it is implemented sequentially. Considering a new variant, we found that depending on the population immunity and transmissibility of the variant, wearing masks and other interventions may be necessary for controlling the disease.

**Funding:** This paper is supported by the Korea National Research Foundation (NRF) grant funded by the Korean government (MEST) (NRF-2021M3E5E308120711). This paper is also supported by the Korea National Research Foundation (NRF) grant funded by the Korean government (MEST) (NRF-2021R1A2C100448711). This research was also supported by a fund (2022-03-008) by Research of Korea Disease Control and Prevention Agency.

**Research in context:** *Evidence before this study:* There are numerous studies in modelling transmission dynamics of COVID-19 variants but only a few published works tackle the lifting of mask mandate considering the omicron variant, although these studies did not consider unreported cases, variants, and waning immunity. Furthermore, there is no age-structured modeling study which investigated the effect of lifting mask mandate considering high immune state of the population, contributed by both of natural infection and vaccination.

*Added value of this study:* Our mathematical model considered key factors such as vaccine status, age structure, medical staff, prior infection, and unreported cases to study the COVID-19 epidemic in Korea. Updated data and variant-specific parameters were used in the model. Contact patterns in the household, school, work, hospital and other places are considered separately to make the model applicable to the mask mandate issue. Seasonality and scenarios on possible future variants are also included in this study.

*Implications of all the available evidence:* With mask wearing as one of the remaining non-pharmaceutical interventions in Korea and other countries, this study proposes strategies for lifting the mask mandates while ensuring that cases remain manageable. A variant-dependent factor is incorporated into the model so that policymakers could prepare proactive intervention policies against future variants.

## Introduction

The Omicron variant has altered the course of the COVID-19 pandemic in Korea. The number of confirmed cases from February 1 to February 18, 2022 already exceeded the number of cumulative confirmed cases before February 1, 2022.^1^ During the first Omicron wave in 2022, the peak number of administered severe patients^*^ reached more than 1,200 in mid-March, while the maximum capacity for the severe patients at the end of March was approximately 2,000.^1,3^ As the number of cases skyrocketed, the number of PCR tests became insufficient. In turn, the Korean government changed the testing policy to include results from rapid antigen tests (RAT) conducted at medical clinics or hospitals. Since RAT has a lower accuracy than PCR and as symptoms became milder, unreported cases in Korea are expected.^4^ Indeed, the Korea Disease Control and Prevention Agency performed antibody tests to random sampled 9,901 people in August 2022 and found that there was a 19·15% difference between the case-confirmed ratio (38·50%) and N-antibody-positive ratio (57·65%), which is specifically induced after natural infection.^5^

Even though the reported cases were high, nonpharmaceutical interventions (NPIs) were gradually eased due to the significantly milder symptoms and nationwide vaccination programs. Social distancing, which has been the primary disease control strategy in Korea before the vaccines and antiviral drugs were available, was lifted on April 18, 2022.^6^ Since then, wearing masks outdoors has become non-compulsory, while the policy on wearing masks indoors has remained in place.^5^ Still, there are potential risks in easing NPIs. For example, at the end of 2021, the Korean government implemented a ‘return to normal’ policy, which eases interventions gradually. However, the number of administered severe patients at that time reached more than 1,100.^7^ Careful consideration of many aspects such as vaccination, variants, booster shots, waning immunity, and age-specific factors is necessary to properly provide guidance to policymakers in decision-making.

Since the COVID-19 outbreak, policymakers turn to mathematicians in predicting cases and crafting strategies that can mitigate the spread of infection.^8,9,10^ There are numerous modelling studies considering the COVID-19 variants.^11–15^ However, only a few published works tackle the lifting of the mask mandate considering the Omicron variant. An evaluation of the different types of face masks has been proposed.^16^ A limitation of their study is that vaccination was not incorporated. Vaccination and mask wearing are studied but the authors assumed that the population is well-mixed and age-specific factors were not considered.^17^ Age-structured models that considers vaccination were proposed.^18,19^ However, unreported cases, booster shots, and specific variants were not considered.

In this study, we developed a model of COVID-19 to analyze the Omicron outbreak in Korea considering vaccination history (primary, booster, and updated booster shots), age structure that distinguishes the medical staff from the general population, breakthrough infections, unreported cases, seasonality, and NPIs. Since indoor mask wearing is the remaining mandatory NPI in Korea, we investigate the effect of lifting this policy in all locations. In the model, we quantify the effects of easing the mask protocol in school, work, other places, and hospital settings using contact rates between the different age groups. The model uses updated variant-specific vaccine effectiveness. Hence, scenarios on possible new variants can be studied. To the best of our knowledge, our work is the only age-structured mathematical model that considers updated booster shots and mask wearing. The proposed model can be applied not only to Korea, but also to other countries that are yet to remove all NPIs. As of December 2022 in the Western Pacific Region, China has most strict mask mandates, which requires to wear masks in outdoor.

## Methods

### Mathematical modeling of COVID-19

In the model, we divided the population according to age, medical staff (MS) or non-MS, with or without prior infection, vaccination, and infection status. The epidemiological process follows a basic SEIR structure, in which the severity of the disease and unreported cases were added. Figure 1 illustrates the flowchart of the mathematical model. There are 12 stages considering vaccine and prior infection: unvaccinated, *u*, primary vaccinated within 180 days, *v*, primary vaccinated after 180 days, 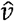, boostered within 180 days, *b*, boostered after 180 days, 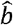, updated booster administrated, 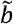. The subscript (*pi*) indicates that the host experienced prior infection. There are five age groups considered: 0-19 (*I*), 20-29 (*II*), 30-49 (*III*), 50-64 (*IV*), and 65+ (*V*). Assuming that there are no underage or senior medical staff, additionally, there are three MS groups: 20-29 MS (*II*_*MS*_), 30-49 MS (*III*_*MS*_), and 50-64 MS (*IV*_*MS*_). The initial time of our model simulation is August 15, 2022, which is in the middle of an outbreak, and therefore, the initial state of the model is not a disease-free state. Details on the setting of the initial states of the model are described in the Supplementary material. We also consider the waning of immunity due to vaccination (*ω*) and the waning of immunity from natural infection (*ζ*). The system of equations describing the model are in the Supplementary material.

**Figure 1.**
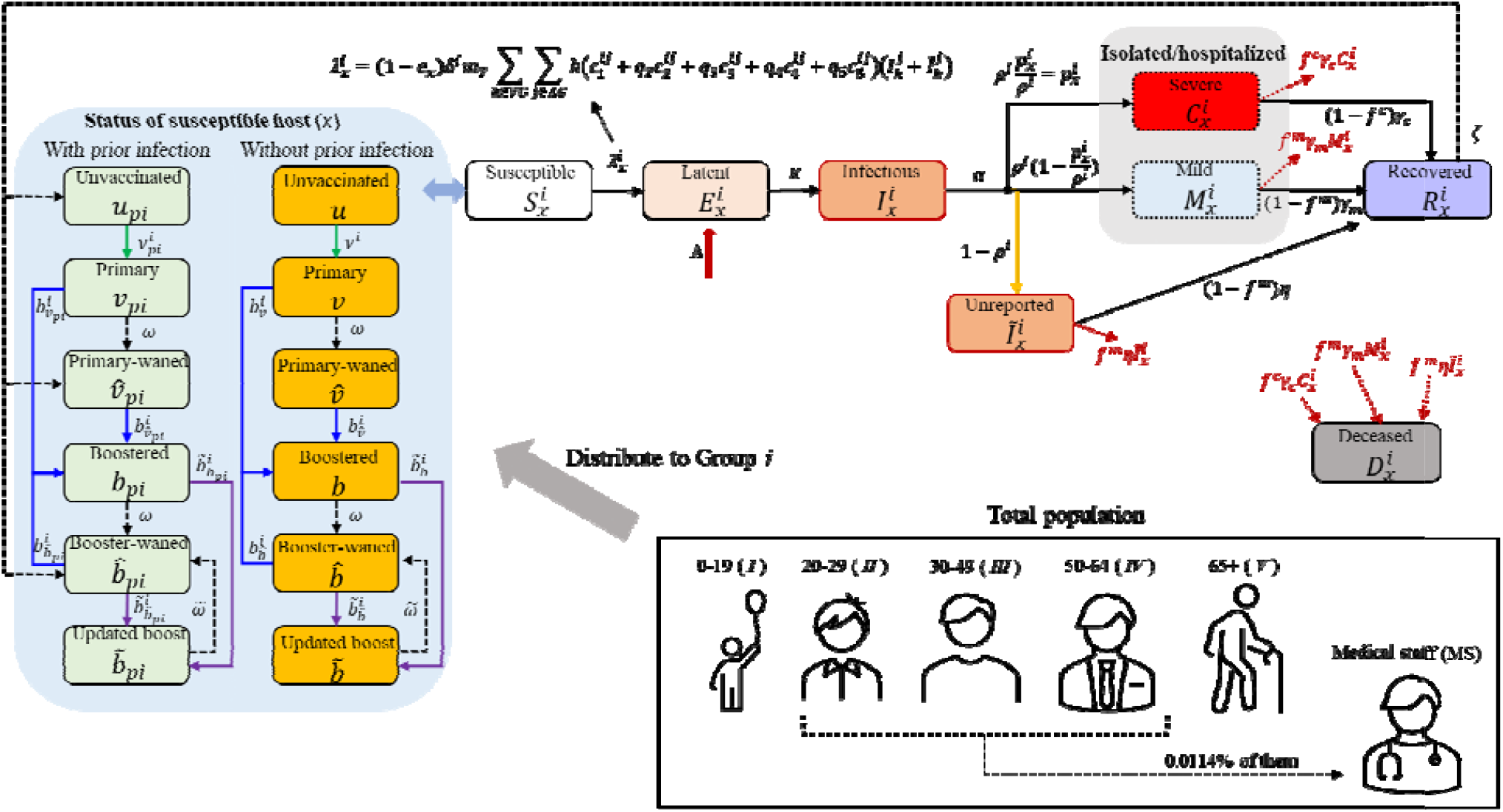
Flow diagram of the mathematical model of COVID-19 outbreak

In Figure 1, the force of infection to the susceptible age group *i* ∈ *AG* in stage *x* ∈ *VG*, denoted as 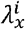, is formulated as

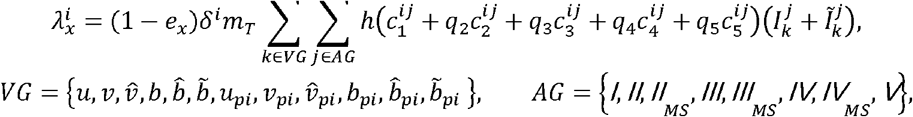

Where 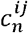 indicates the average number of contacts from age group *i* to *j*, and the subscript *n* indicates the type of contact: 1 for household, 2 for school, 3 for work, 4 for other places, and 5 for hospital. The contact patterns *c*_1_ to *c*_4_ were aggregated and adjusted according to the age structure of our model, while *c*_5_ was estimated assuming that one medical doctor and nurse (2 medical doctors and 3 nurses) have contact with an outpatient (inpatient) per day in Korea.^20–23^ The age-dependent relative susceptibility compared to age 0-19 (*δ*^*i*^, i.e.,*δ*^*I*^ = 1) takes into account clinical susceptibility and other factors such as compliance to policy or behavior. The parameter *q*_*n*_ (*n*: 2, 3, 4, 5) indicates the relative risk of contact from type *n* compared to the household contact 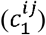, *h* is the probability of disease transmission through household contacts, and *m*_*T*_ is the relative seasonality factor compared to the initial time, i.e.,. Eight parameters, *h, δ*^*II*^, *δ*^*III*^, *δ*^*IV*^, *δ*^*V*^, *m*_2_, *m*_3_, and *m*_4_, are estimated using the Metropolis-Hastings algorithm, which is Markov chain Monte Carlo method to estimate the distribution of unknown parameters, to fit the cumulative confirmed cases of five age groups from August 15 to November 30, 2022 (108 days).^1,24^

### Scenario setting

In this study, we considered three phases (illustrated in Figure 2). In the first phase (dark arrow), we estimated the model parameters using age-dependent cumulative confirmed case data from August 15, to November 30, 2022.^1^ Then we extended our model simulation for 180 days to observe the impact of lifting the mask mandate (blue arrow) on the number of cases. For the extension, we used the estimated relative seasonality factor of the last phase (*m*_4_). We set that if mask-wearing becomes non-mandatory in a certain location, then the transmission risk in that location (*q*_1_ to *q*_5_) increases 5-fold.^25^ Finally, the simulation is extended further for another 180 days (orange arrow) to consider the effect of a new variant. We assume that the new variant has higher transmissibility and risk of breakthrough infections. We incorporate both risks as a single parameter, which we refer to as the variant varying factor (*VVF*), which reduces vaccine effectiveness and increases transmissibility in the force of infection for the forecast 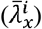 as follows,

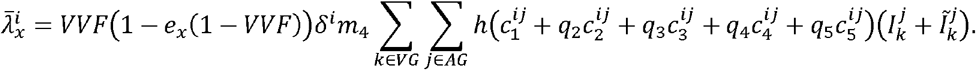

**Figure 2.**
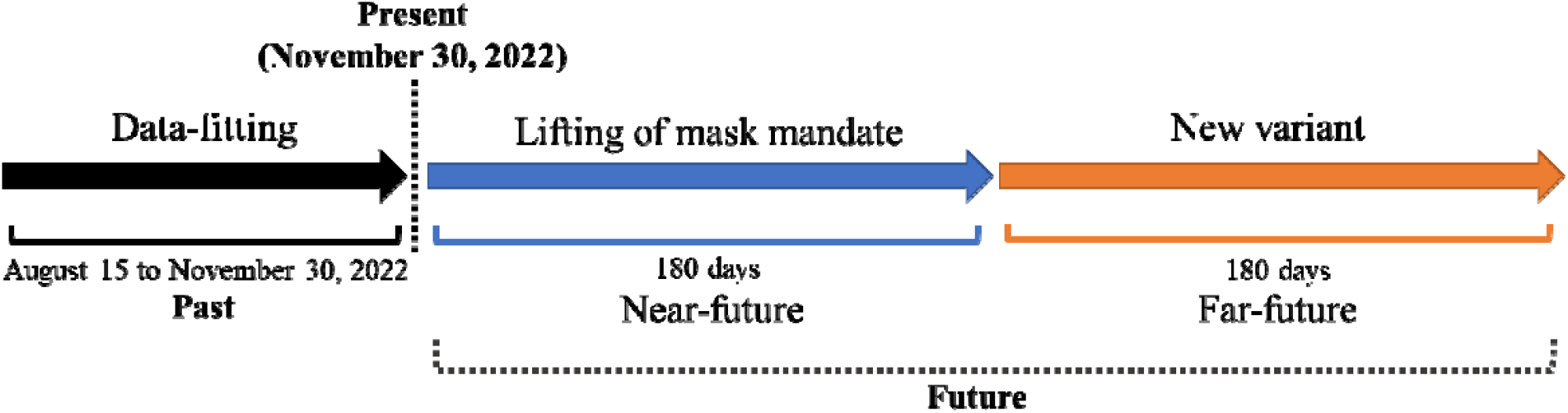
Timeline of the study considering three phases.

In the analysis considering *VVF*, we focused on the peak size of the administered severe patients and set the critical number at 2,000, which is nearby the maximum severe patient capacity in 2022.^1,3^ We also assumed that updated vaccines are administered as the new variant emerges and have relatively higher vaccine effectiveness (if the host does not have prior infection, then 0·8 against infection and 0·95 against severity, if does, then 0·9 against infection and 0·95 against severity) during this phase. Note that we did not consider reduction of vaccine effectiveness against severity caused by new variant.

## Results

### Data-fitting result

The mode of the obtained samples for *h, δ*^*II*^, *δ*^*III*^, *δ*^*IV*^, *δ*^*V*^, *m*_2_, *m*_3_, and *m*_4_ using the Metropolis-Hastings algorithm were used in the model. The corresponding values are 0·52, 1·72, 0·97, 1·53, 2·75, 0·93, 1·51, and 1·26, respectively. Figure 3 shows the aggregated contact matrices and formulated transmission matrix among age groups using the estimated values. The transmission matrix in panel (f) shows distinctive non-symmetric shape as it contains the age-dependent relative susceptibility (*δ*^*i*^)e.g., 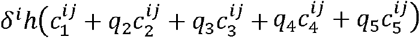.

**Figure 3.**
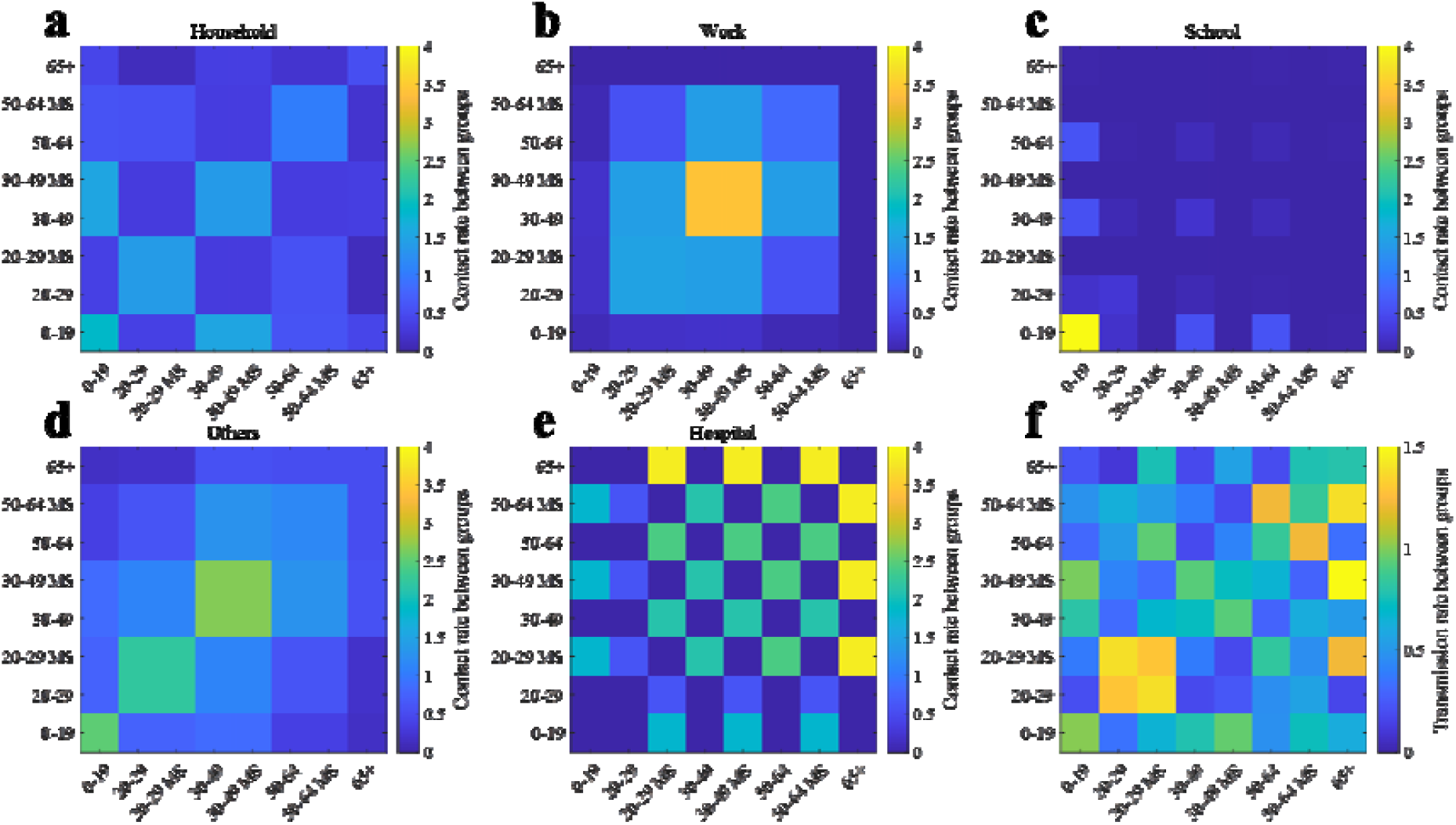
Matrices used in the mathematical model. (a) to (e) the contact rate among age groups. (f) Transmission rates among age groups using matrices (a) to (e).

The fitted curves for the five age groups are displayed in Figure 4. Note that the MS results are included in their corresponding age groups. Panel (a) of Figure 5 shows the effective reproductive number (*R*_*t*_) calculated using the next-generation method.^26^ The filled areas indicate the age-related proportion 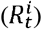, which is computed by summing the columns of the next-generation matrix.^26^ The formulation of the next-generation matrix is described in the Supplementary material. Moreover, panels (b) and (c) show the pie charts that depict the proportion of the population and the average contribution of 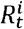 to *R*_*t*_, respectively. The relative difference between population proportion and dedication to reproductive number of age 0-19, 20-29, 30-49, 50-64, and 65+ were 5·01%p, −13·36%p, 0·10%p, 8·88%p, and 0·63%p, respectively, where the negative sign indicates that the group dedicates the transmission more than their proportion to the population.

**Figure 4.**
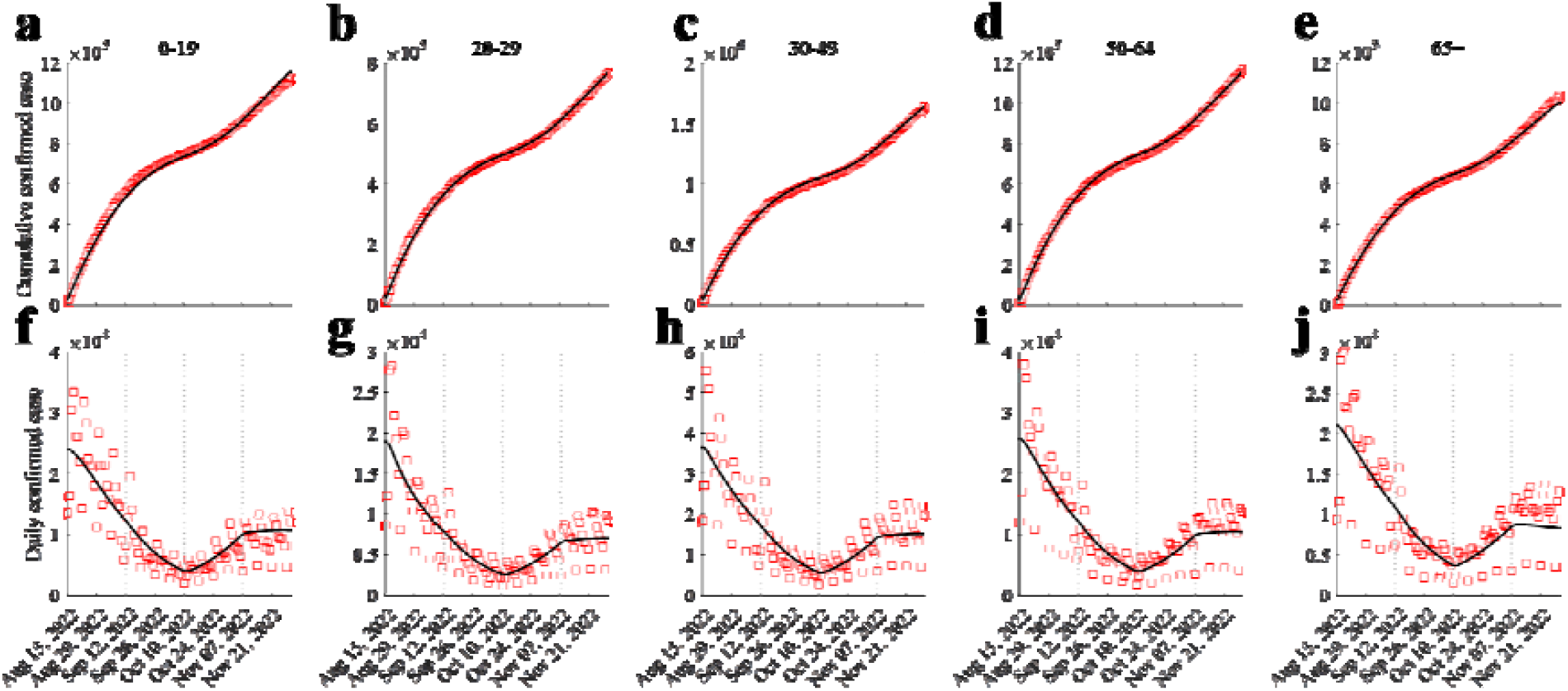
Data-fitting simulation results. Red squares and dark curves indicate real data model and simulation results, respectively.

**Figure 5.**
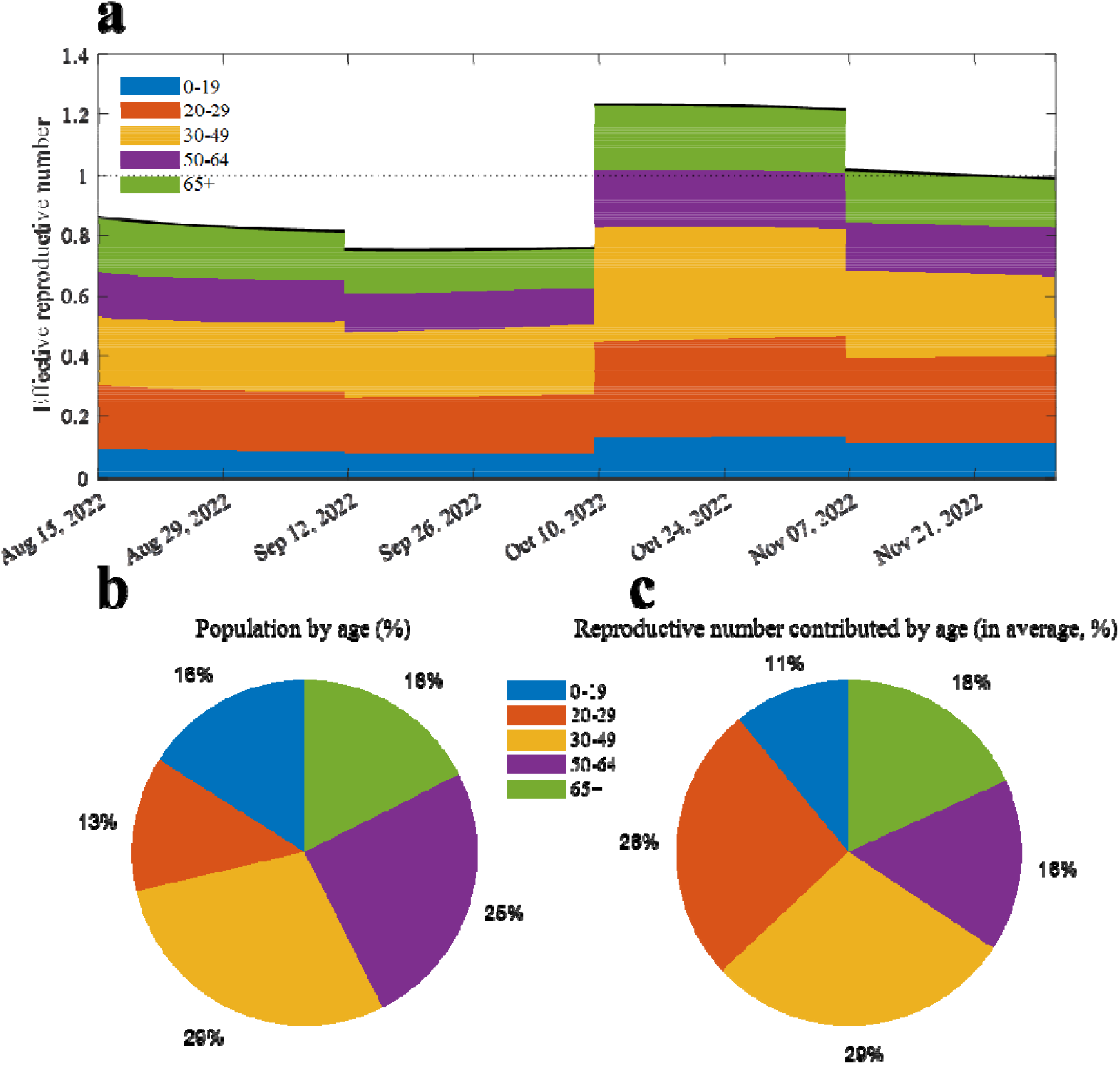
Reproductive number from the model simulation. (a) Effective reproductive number (dark curve), filled area indicates how much each age dedicate to the value. (b) Proportion of the population by age. (c) Average proportion of the reproductive number contributed by age.

### Near-future forecast with lifted mask mandate

Figure 6 shows the simulation results considering the lifting of the mask mandate in various places. Panels (a) and (b) ((c) and (d)) display the daily and cumulative confirmed cases (administered severe patients), respectively. Excluding the masks-on scenario, removing masks in schools resulted in both the lowest number of cumulative confirmed cases (7,501,510) and severe patients (71,710). Taking off masks everywhere and except in hospitals have the first and second highest cumulative number of severe patients, respectively. Lifting of mask mandates in hospitals showed the highest number of confirmed cases and severe patients among the lifting mask mandate in single place settings (set.1 to set.4). Adding the medical staffs to their respective age groups, the confirmed cases (severe patients) in ages 0-19, 20-29, 30-49, 50-64, and 65+ in every scenario were approximately 19·59% (00·16%), 13·48% (00·80%), 29·02% (09·92%), 20·16% (37·22%), and 17·75% (51·90%), respectively. The effective reproductive numbers in each scenario are shown in panel (e). The initial value of the reproductive number for scenarios 1 to 8 were ranged from 0·99 to 1·19. Panel (f) shows the age-dependent increment in the reproductive number in scenarios 1 to 3. If masks are taken off in school, the underage group has the highest increment (3·24%) of their reproductive number while the increments in other age groups were less than 1%.

**Figure 6.**
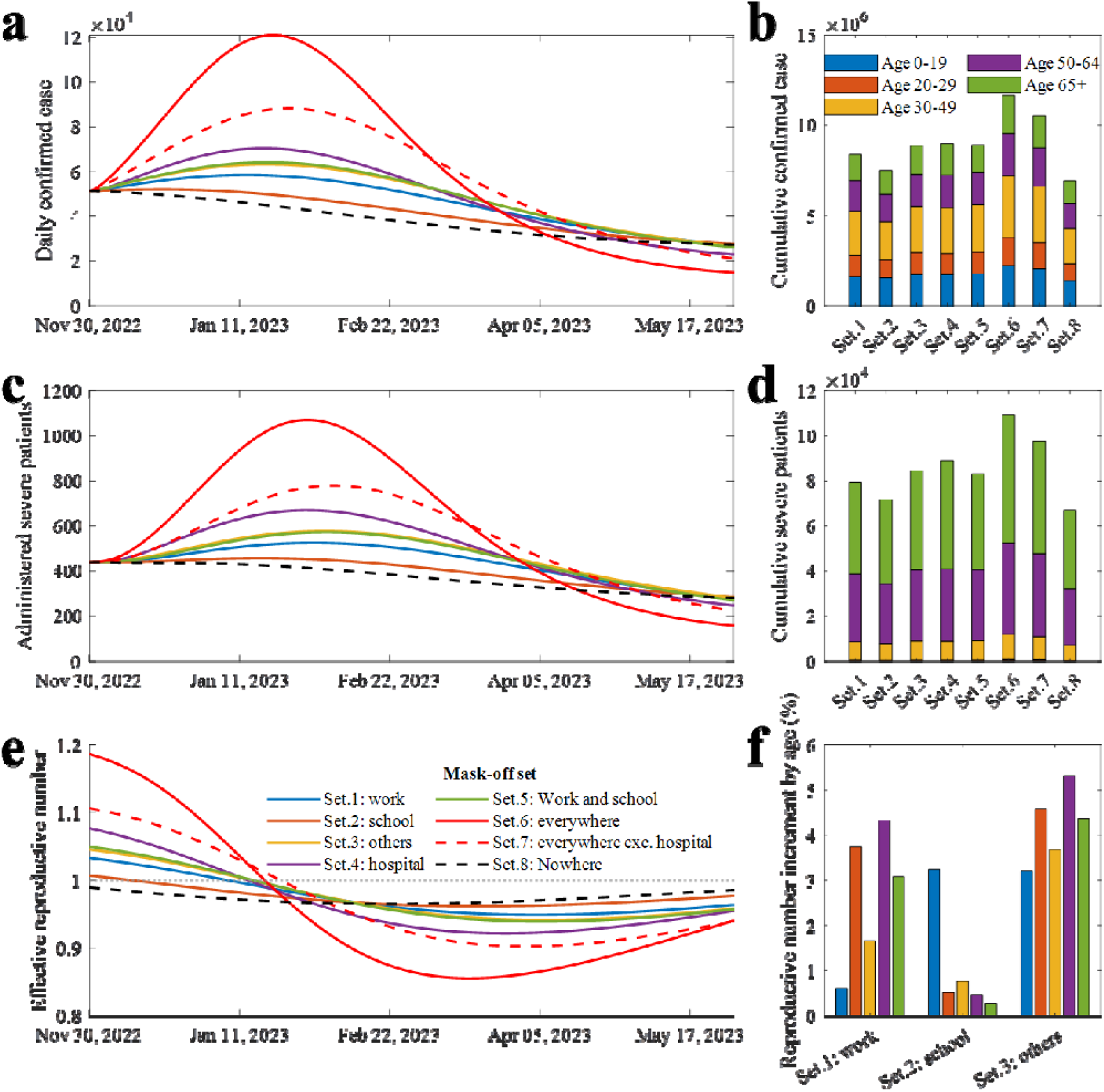
Extended simulation results considering lifted mask mandate. (a) Daily confirmed cases. (b) Cumulative confirmed cases of age groups in different scenarios. (c) Administered severe patients. (d) Cumulative severe patients of age groups in scenarios. (e) Effective reproductive number. (f) Age dependent transmissibility increment in different scenarios.

Later, we investigated the effect on the number of cases of a sequential lifting of the mask mandate in different locations (Figure 7). In the simulations, we set that mask wearing in hospitals is mandatory. Lifting the policy in school, followed by the workplace, and then in other places (school-work-other) showed the least number of confirmed cases and severe patients, while other-work-school had the greatest number of confirmed and severe patients. The other-work-school sequence resulted in 6·94% (7·89%) higher confirmed cases (severe patients) than when the order is school-work-other.

**Figure 7.**
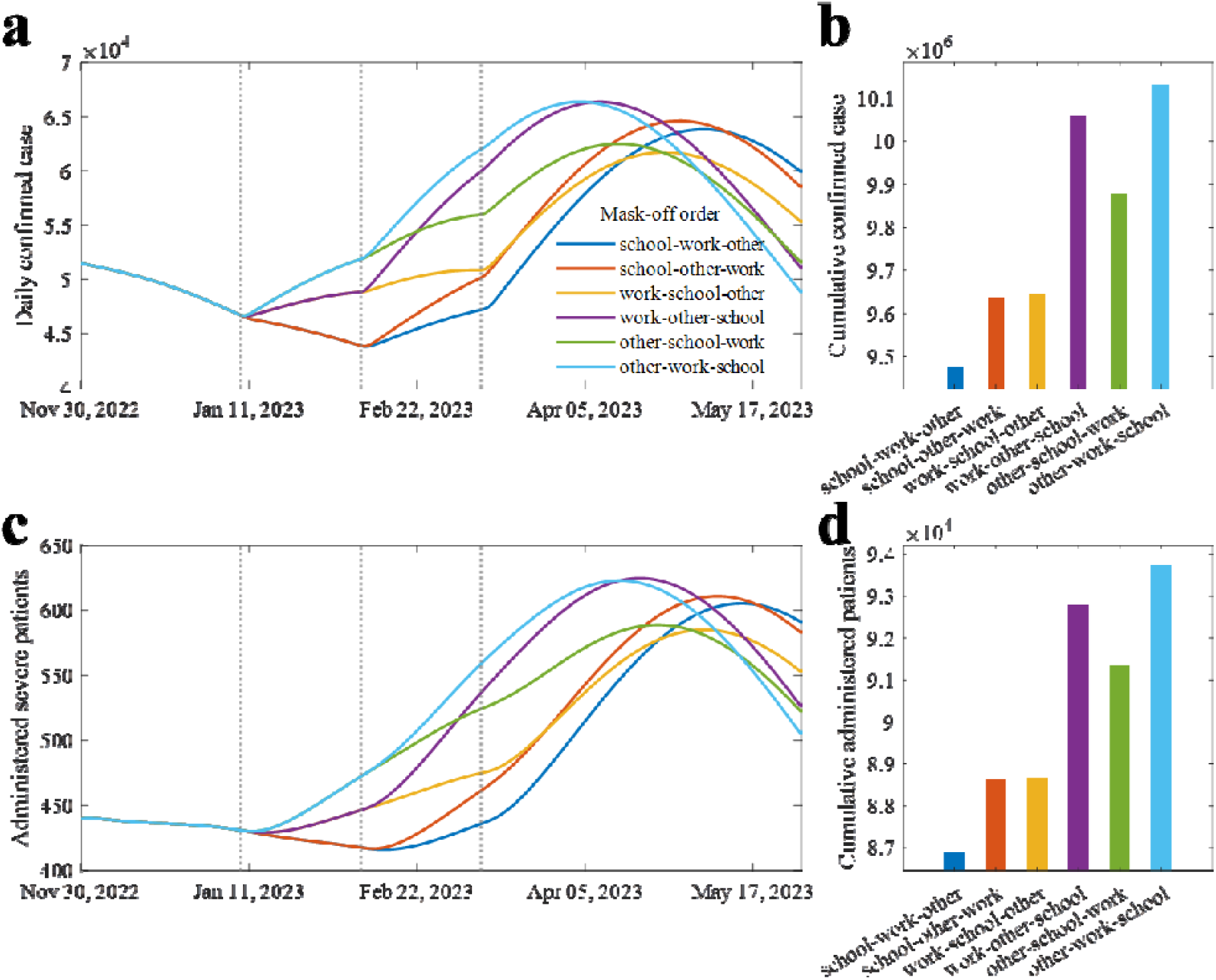
Extended simulation results with sequentially lifted mask mandate. (a) Daily confirmed cases. (b) Cumulative confirmed cases in scenarios. (c) Administered severe patients. (d) Cumulative severe patients in scenarios.

### Far-future forecast with new variant

We extended the model simulation for 180 days from November 30, 2022 to forecast the number of cases considering the possible emergence of a new variant. We illustrate these in Figure 8. Assuming different mask-wearing policy, panels (a) to (c) display the heatmaps of the peak number of severe patients depending on *VVF* and number of daily vaccinations. Contour lines are added to indicate critical level of severe cases (2,000). Panel (d) shows the contour lines with three distinctively filled area; (1) best-case scenario (blue region), which does not require mask-wearing, (2) status-quo (green region), which requires hybrid mask-wearing, that is, masks may not be required in certain places, and (3) worst-case scenario (yellow area), which means the peak number of severe patients exceeds the critical level. Panel (e) and (f) illustrate sampled simulation results, from panel (a) to (c) to visualize the forecasted number of cases over time. If *VVF* = 0·35, severe cases may reach more than 2,000, which means additional NPIs is required. On the other hand, if *VVF* = 0·2, the number of cases remains manageable even if the mask mandate is removed.

**Figure 8.**
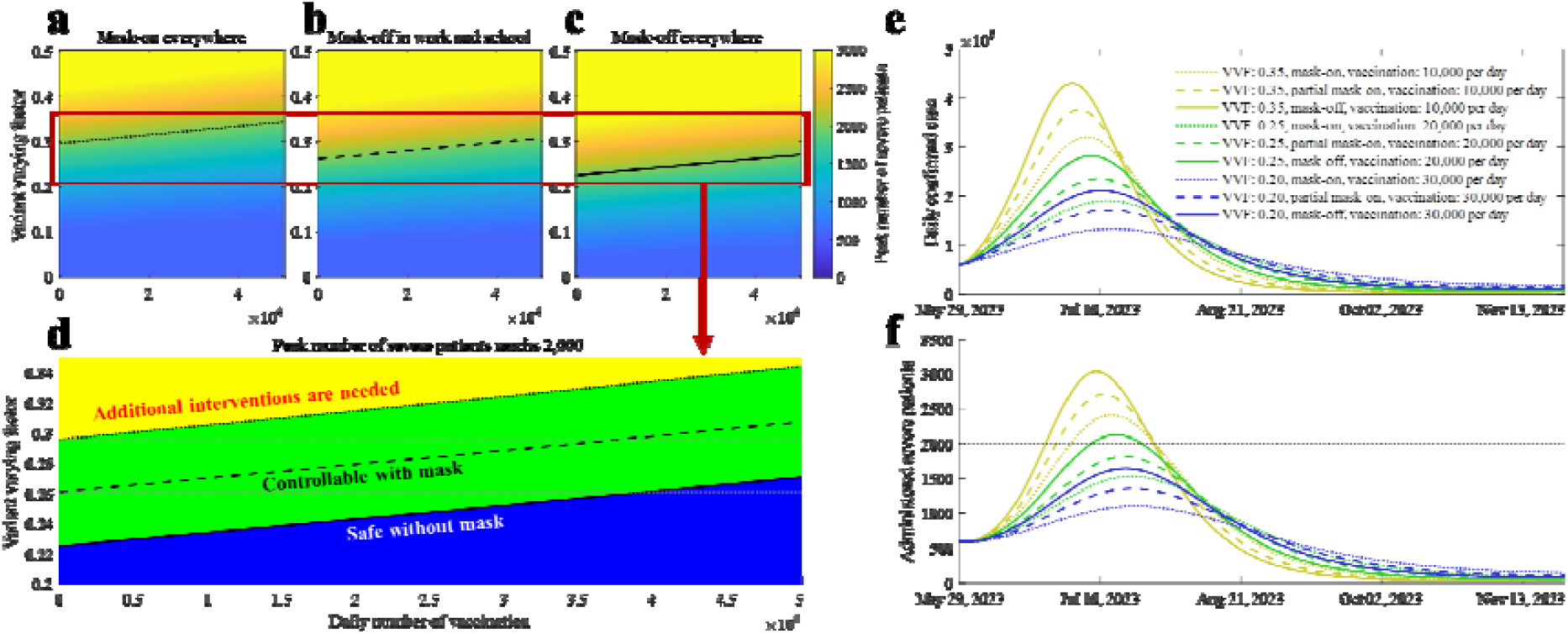
Peak size of administered severe patients in different settings. (a) When there is mask mandate in everywhere. (b) When mask mandate is lifted in work and school. (c) When the mask mandate is lifted in everywhere except hospital. (d) Separable three areas using the contour lines from panel (a) to (c). Note that contour lines in (a) to (d) indicate the peak size of administered severe patient as 2,000. (e) to (f) show the simulation results sampled from (a) to (c).

## Discussions

Our proposed model considers different key factors to make it more realistic. A complex model that is not parameterized well might be less useful in rendering the natural phenomena compared to a simple model.^27^ Although our proposed model requires estimation of several parameters, we made sure to aggregate the most updated data. The most challenging part in our estimation is the setting of the initial state values, because we initialize our simulation in the middle of the pandemic, which necessitates information about prior infections. Fortunately, this is possible in this study as antibody test data are available.

Results of the Metropolis-Hastings algorithm provide estimates as well as uncertainties of the parameter estimates. A low prior infection ratio of seniors and a high susceptibility to infection shown by the model pose potential risks in the next wave. Moreover, a high contact rate between seniors and MS can be seen, which suggests that removing mask protocols in hospitals is not advisable given that the number of severe cases is still high.^28^

On the other hand, a high prior infection proportion for those aged below 20 resulted to reduced infectivity. This suggests that wearing masks inside schools has less effect in reducing cases in current situation, although it was known as highly effective measure before omicron.^29^ The transmissibility of the disease is reflected in the estimated relative seasonality factors (*m*_*T*_), as it decreased from the middle of August to early October but later on increased. This change can be attributed to the characteristics of the virus and the seasonal social behavior, for example, more outdoor activities during summer months and more indoor activities during colder months. In Korea, since wearing masks indoors is the only remaining NPIs, the natural seasonality of the disease can be investigated. We expect that by incorporating this into the model, a more dynamic and realistic policy can be formulated to cope with the pandemic.

Most of the severe cases were observed to have occurred in the older age groups (in Figure 6). Since seniors have more frequent contact with MS than the other age groups, our model simulations highlight the importance of mask wearing in hospitals. On the other hand, taking off masks in schools might be acceptable. We already observed that the underage group has highest prior infection (73·40%, described in the Supplementary material) and low reproductive number (in Figure 5). Moreover, the increment in the reproductive number in the underage is small and affects the other age groups minimally (panel (f) in Figure 6). The lifting of the mask mandates is possible and may be more manageable if the policy is relaxed sequentially. We found that if masks are removed in all locations at once except in hospitals, then the administered severe patients would reach 800. On the other hand, if the policy is relaxed in a sequence (school-work-other), then the severe patients are below 650 and decrease further.

If a highly transmissible new variant emerges, a risk of health system saturation is possible because of the high number of severe patients. We observed three possibilities depending on the VVF, vaccination rate, and threshold of severe patients. The critical *VVF* value is approximately 0·3. Mask wearing may still be necessary if the *VVF* is higher than 0·23. For comparison, assuming that the population has 0·8 of the protection effect against infection, then the effective reproductive number would become 3·24 folds if new variant has 0·35 of *VVF*, i.e. [(1.35 × (1 − 0.8 × (1 − 0.35))} ÷ {1 × (1− 0.8)}] = 3· 24. It was found that the relative reproduction number of omicron compared to delta was 3·19, which would be similar to the previous example when *VVF* was assumed as 0·35.^30^ The *VVF* proposed in this study can help policymakers in coming up with proactive intervention strategy when a new variant arises. When a more contagious variant becomes dominant, vaccination and mask-wearing may still both be essential to reduce the number of severe cases. For example, if the number of daily-administered vaccines is 5,000 (approximately 0.01% of the population) and *VVF* is 0·32, then mask-wearing may not be enough. On the other hand, the outbreak may be controllable with mask-wearing if the number of daily-administered vaccines is 40,000. As the scale of vaccination increases, the disease becomes more manageable even when *VVF* is high. Thus, efforts to reduce vaccine hesitancy in the older age group should be implemented as this group has the highest proportion of severe cases.

### Limitation and future study

In this study, we assumed that the N-positive proportion is the proportion of hosts who had prior infection. As with other serological tests, the N-positive immune response may not be detected if the infection was much earlier, which means that the actual proportion of population with prior infection would be higher than the investigation results. Furthermore, we assumed that N-positive rate of age 0-4 is the same as that of age 5-9. In the model parameter setting, the effectiveness of updated vaccine is assumed due to the lack of study. In initial states, the proportion of waned infection-induced immunity is set to 50%. We recognize that the contact matrix, N-positive rate for age 0-4, and effectiveness of updated vaccines can still be properly estimated but these can easily be modified in the model once the data become available.

## Conclusion

In this study, we analyzed the risks related to lifting the mask mandate and new variants using a well-structured mathematical model. We found that removing the mask policy is possible while keeping the number of cases manageable. Notably, results show that masks off in school can be implemented sooner. However, masks in hospitals should remain in place to protect the older age groups. Possible new variants might cause the rise in infections and require stricter NPIs if the new variant has 30% higher transmissibility and immune reduction compared to the current omicron variant.

Simulations showed a hopeful perspective as we observed that the critical variant varying factor is about 0·3, which was similarly recorded once in the past, when delta was replaced by omicron as the dominant variant, with the severity of omicron is significantly lesser than delta. Still, the potential risk is never zero and prompt characterization of the new variant is vital as it can guide government to decide the proactive response. Although the model is formulated using Korean data, it is general enough that it can be applied to other countries who are yet to lift all their COVID-19 restrictions.

## Data Availability

All data produced are available online

## Acknowledgment

This paper is supported by the Korea National Research Foundation (NRF) grant funded by the Korean government (MEST) (NRF-2021M3E5E308120711). This paper is also supported by the Korea National Research Foundation (NRF) grant funded by the Korean government (MEST) (NRF-2021R1A2C100448711). This research was also supported by a fund (2022-03-008) by Research of Korea Disease Control and Prevention Agency.

We appreciate the Korean Society for Industrial and Applied Mathematics for organizing the thematic program ‘KSIAM-NIMS School on Biomathematics’.

