## Supplementary appendix for "Can Koreans be ‘FREE’ from mask wearing?: Advanced mathematical model can suggest the idea"

**System equations of the mathematical model of COVID-19 outbreaks in Korea**

The system of equations describing our model is as follows,

$$\frac{dS_{x}^{i}}{dt}=-\lambda_{x}^{i}S_{x}^{i}+IN_{x}^{i}-OUT_{x}^{i},$$

$$\frac{dE_{x}^{i}}{dt}=\lambda_{x}^{i}S_{x}^{i}+\Lambda-\kappa E_{x}^{i},$$

$$\frac{dI_{x}^{i}}{dt}=\kappa E_{x}^{i}-\alpha I_{x}^{i},$$

$$\frac{dM_{x}^{i}}{dt}=\alpha\rho^{i}\left( 1-\frac{\left( 1-\bar{e}_{x} \right)p^{i}}{\rho^{i}} \right)I_{x}^{i}-\gamma_{m}M_{x}^{i},$$

$$\frac{dC_{x}^{i}}{dt}=\alpha\left( 1-\bar{e}_{x} \right)p^{i}I_{x}^{i}-\gamma_{c}C_{x}^{i},$$

$$\frac{d\tilde{I}_{x}^{i}}{dt}=\alpha\left( 1-\rho^{i} \right)I_{x}^{i}-\eta\tilde{I}_{x}^{i},$$

$$\frac{dR_{x}^{i}}{dt}=\left( 1-f_{m} \right)\gamma_{m}M_{x}^{i}+\left( 1-f_{c} \right)\gamma_{c}C_{x}^{i}+\left( 1-f_{m} \right)\eta\tilde{I}_{x}^{i}-\zeta,$$

$$\frac{dD_{x}^{i}}{dt}=f_{m}\gamma_{m}M_{x}^{i}+f_{c}\gamma_{c}C_{x}^{i}+f_{m}\eta\tilde{I}_{x}^{i},$$

$$\lambda_{x}^{i}=\left( 1-e_{x} \right)\delta^{i}m_{T}\sum_{k\in VG} \sum_{j\in AG} h\left( c_{1}^{ij}+q_{2}c_{2}^{ij}+q_{3}c_{3}^{ij}+q_{4}c_{4}^{ij}+q_{5}c_{5}^{ij} \right)\left( {I_{k}^{j}+\tilde{I}}_{k}^{j} \right),$$

$$IN_{u}^{i}=0, {OUT}_{u}^{i}=\nu^{i},$$

$$IN_{v}^{i}=\nu^{i}, {OUT}_{v}^{i}=\omega S_{v}^{i}+b_{v}^{i},$$

$$IN_{\hat{v}}^{i}=\omega S_{v}^{i}, {OUT}_{\hat{v}}^{i}=b_{\hat{v}}^{i},$$

$$IN_{b}^{i}=b_{v}^{i}+b_{\hat{v}}^{i}+b_{\hat{b}}^{i}, {OUT}_{b}^{i}=\omega S_{b}^{i}+\tilde{b}_{b}^{i},$$

$$IN_{\hat{b}}^{i}=\omega S_{b}^{i}+\tilde{\omega}S_{\tilde{b}}^{i}, {OUT}_{\hat{b}}^{i}=b_{\hat{b}}^{i}+\tilde{b}_{\hat{b}}^{i},$$

$$IN_{\tilde{b}}^{i}=\tilde{b}_{b}^{i}+\tilde{b}_{\hat{b}}^{i}, {OUT}_{\tilde{b}}^{i}=\tilde{\omega}S_{\tilde{b}}^{i},$$

$$IN_{u_{pi}}^{i}=\zeta\left( R_{u}^{i}+R_{u_{pi}}^{i} \right), {OUT}_{u_{pi}}^{i}=\nu_{pi}^{i},$$

$$IN_{v_{pi}}^{i}=\nu_{pi}^{i}, {OUT}_{v_{pi}}^{i}=\omega S_{v_{pi}}^{i}+b_{v_{pi}}^{i},$$

$$IN_{\hat{v}_{pi}}^{i}=\zeta\left( R_{v}^{i}+R_{\hat{v}}^{i}+R_{v_{pi}}^{i}+R_{\hat{v}_{pi}}^{i} \right)+\omega S_{v_{pi}}^{i}, {OUT}_{\hat{v}_{pi}}^{i}=b_{\hat{v}_{pi}}^{i},$$

$$IN_{b_{pi}}^{i}=b_{v_{pi}}^{i}+b_{\hat{v}_{pi}}^{i}+b_{\hat{b}_{pi}}^{i}, {OUT}_{b_{pi}}^{i}=\omega S_{b_{pi}}^{i}+\tilde{b}_{b_{pi}}^{i},$$

$$IN_{\hat{b}_{pi}}^{i}=\zeta\left( R_{b}^{i}+R_{\hat{b}}^{i}+R_{\tilde{b}}^{i}+R_{b_{pi}}^{i}+R_{\hat{b}_{pi}}^{i}+R_{\tilde{b}_{pi}}^{i} \right)+\omega S_{b_{pi}}^{i}+\tilde{\omega}S_{\tilde{b}_{pi}}^{i}, {OUT}_{\hat{b}_{pi}}^{i}=b_{\hat{b}_{pi}}^{i}+\tilde{b}_{\hat{b}_{pi}}^{i},$$

$$IN_{\tilde{b}_{pi}}^{i}=\tilde{b}_{b_{pi}}^{i}+\tilde{b}_{\hat{b}_{pi}}^{i}, {OUT}_{\tilde{b}_{pi}}^{i}=\tilde{\omega}S_{\tilde{b}_{pi}}^{i},$$

where $x∊VG=\{u,v,\hat{v},b,\hat{b},\tilde{b},u_{pi},v_{pi},\hat{v}_{pi},b_{pi},\hat{b}_{pi},\tilde{b}_{pi}\}$, $i∊AG=\{Ⅰ,Ⅱ,Ⅱ_{MS},Ⅲ,Ⅲ_{MS},Ⅳ,Ⅳ_{MS},Ⅴ\}$, and vaccine administration per day parameters, {$\nu^{i}$, $b_{v}^{i}$, $b_{\hat{v}}^{i}$, $b_{\hat{b}}^{i}$, $\tilde{b}_{b}^{i}$, $\tilde{b}_{\hat{b}}^{i}$, $\nu_{pi}^{i}$, $b_{v_{pi}}^{i}$, $b_{\hat{v}_{pi}}^{i}$, $b_{\hat{b}_{pi}}^{i}$, $\tilde{b}_{b_{pi}}^{i}$, $\tilde{b}_{\hat{b}_{pi}}^{i}$}, correspond to the data on daily number of administered vaccines.^1,2^ The model parameters are listed in Supplementary Table 1. In our model, two types of vaccine effectiveness are used: against infection ($e_{x}$) and against severity ($\bar{e}_{x}$). The values were adjusted according to the effectiveness and proportion of an age group vaccinated with the different types of vaccines administered in Korea. The details of the calculation are described in our past research.^3^ For hosts with prior infection, we assumed that the waned-vaccine effectiveness is the same as that of the unvaccinated but infected ($e_{u_{pi}}$, $\bar{e}_{u_{pi}}$). We also assumed that the effectiveness of the updated vaccines against infection and severity are 0·8 and 0·95 (0·8 and 0·95), respectively, if the host does not have a prior infection (host has prior infection). In Korea, updated vaccines have been administered since October 11, 2022.^4^ Supplementary Table 2 shows the values for the vaccine effectiveness that were used in this study.

**Supplementary Table** 1 Model parameters

| **Symbol** | **Description** | **Value** | **Reference** |
| --- | --- | --- | --- |
| $q_{2}$ | Relative risk of work contact compared to household contact | 0·01 | ^5^ |
| $q_{3}$ | Relative risk of school contact compared to household contact | 0·01 | ^5^ |
| $q_{4}$ | Relative risk of other contact compared to household contact | 0·01 | ^5^ |
| $q_{5}$ | Relative risk of hospital contact compared to household contact | 0·19 | ^5^ |
| $1/\kappa$ | Latent period | 2 (days) | ^6,7,8,9^ |
| $1/\alpha$ | Infectious period of reported case | 4 (days) | ^9,10^ |
| $1/\eta$ | Extra infectious period of unreported case | 2 (days) | ^11^ |
| $\Lambda$ | Overseas entrant case per day | 1·9479 | ^12^ |
| $\rho^{i}$ | Report rate | 85·6867% (age 0-19)  76·2791% (age 20-29)  73·2376% (age 30-49)  54·4158% (age 50-64)  68·9598% (age 65+) | ^13^ |
| $p^{i}$ | Case severe rate | 0·0018% (age 0-19)  0·0158% (age 20-29)  0·0735% (age 30-49)  0·2977% (age 50-64)  0·6607% (age 65+) | ^12,14^ |
| $\gamma_{c}$ | Recovery period of severe case | 10 (days) | ^15^ |
| $\gamma_{m}$ | Recovery period of mild case | 7 (days) | ^16^ |
| $f_{c}$ | Case fatality rate of severe case | 28·55% | ^17^ |
| $f_{m}$ | Case fatality rate of mild case | 0·0070% | ^12^ |
| $1/\omega$ | Waning period of vaccine-induced immunity | 180 (days) | ^18^ |
| $1/\tilde{\omega}$ | Waning period of updated vaccine-induced immunity | 180 (days) | Assumed |
| $1/\zeta$ | Waning period of infection-induced immunity | 360 (days) | ^18^ |

**Supplementary Table** 2 Vaccine effectiveness

| **VE against** | **Prior infection** | **Phase** | **Symbol** | **Value** | **Reference** |
| --- | --- | --- | --- | --- | --- |
| Infection | No | Primary | $e_{v}$ | 0·37 | ^19,20^ |
|  |  | Primary-waned | $e_{\hat{v}}$ | 0·12 | ^19,20^ |
|  |  | Booster | $e_{b}$ | 0·60 | ^19,20^ |
|  |  | Booster-waned | $e_{\hat{b}}$ | 0·13 | ^19,20^ |
|  |  | Updated booster | $e_{\tilde{b}}$ | 0·8 | Assumed |
|  | Yes | Unvaccinated | $e_{u_{pi}}$ | 0·52 | ^18^ |
|  |  | Primary | $e_{v_{pi}}$ | 0·55 | ^18^ |
|  |  | Primary-waned | $e_{\hat{v}_{pi}}$ | 0·52 | Assumed |
|  |  | Booster | $e_{b_{pi}}$ | 0·77 | ^18^ |
|  |  | Booster-waned | $e_{\hat{b}_{pi}}$ | 0·52 | Assumed |
|  |  | Updated booster | $e_{\tilde{b}_{pi}}$ | 0·8 | Assumed |
| Severity | No | Primary | $\bar{e}_{v}$ | 0·77 | ^21^ |
|  |  | Primary-waned | $\bar{e}_{\hat{v}}$ | 0·45 | ^21^ |
|  |  | Booster | $\bar{e}_{b}$ | 0·90 | ^21^ |
|  |  | Booster-waned | $\bar{e}_{\hat{b}}$ | 0·58 | ^21^ |
|  |  | Updated booster | $\bar{e}_{\tilde{b}}$ | 0·95 | Assumed |
|  | Yes | Unvaccinated | $\bar{e}_{u_{pi}}$ | 0·81 | ^22^ |
|  |  | Primary | $\bar{e}_{v_{pi}}$ | 0·86 | ^22^ |
|  |  | Primary-waned | $\bar{e}_{\hat{v}_{pi}}$ | 0·81 | Assumed |
|  |  | Booster | $\bar{e}_{b_{pi}}$ | 0·94 | ^22^ |
|  |  | Booster-waned | $\bar{e}_{\hat{b}_{pi}}$ | 0·81 | Assumed |
|  |  | Updated booster | $\bar{e}_{\tilde{b}_{pi}}$ | 0·95 | Assumed |

**Initial condition estimation**

The process of dividing the population is illustrated in Supplementary Figure 1. We divide the susceptible population in Korea considering variety of factors; Age, medical staff (MS), vaccine history, and the prior infection. Firstly, entire population is divided into 8 subgroups considering the host’s age and whether the host is medical staff or not ; 0-19 ($Ⅰ$), 20-29 ($Ⅱ$), 30-49 ($Ⅲ$), 50-64 ($Ⅳ$), 65+ ($Ⅴ$) 20-29 MS ($Ⅱ_{MS}$), 30-49 MS ($Ⅲ_{MS}$), and 50-64 MS ($Ⅳ_{MS}$), assuming that there is no underage or senior MS.^23,24^ Later, considering vaccination status in August 15, 2022, divided population is once more divided; unvaccinated, $u$, primary vaccinated within 180 days, $v$, primary vaccinated after 180 days, $\hat{v}$, boostered within 180 days, $b$, boostered after 180 days, $\hat{b}$, updated booster administrated, $\tilde{b}$.^1,2^ Considering high vaccination rate of MS, we set that 90% of MS are in boostered state (45% $b$, 45% $\hat{b}$) and 10% are in $u$. Still, we need to consider proportion of population who experienced infection. We adjusted the proportion considering population size, and set report rate as $\rho^{i}$, which is calculated using the difference between number of reported proportion and actual N-positive proportion. ^13^ Since the investigation did not target age 0-4, we simply set that age 0-4 has same N-positive ratio with age 5-9. By using the N-positive ratio, age-vaccine-subgrouped population is once more subgrouped considering prior infection (subscript $pi$). Proportion of hosts who have prior infection in group $Ⅰ$, $Ⅱ$, $Ⅲ$, $Ⅳ$, and $Ⅴ$ are 73·40, 64·58, 59·68, 54·11, and 43·69, respectively. In this case, vaccine effectiveness was considered to give more proportion of N-positive ratio to unvaccinated hosts. Since most of the COVID-19 infection in Korea occurred in 2022, we simply set that half of hosts with prior infection still remains in $R_{x}^{i}$. Initial state values are listed in Supplementary Table 3.

Since the outbreak situation in mid-August was relatively stable, we simply estimated the initial states of $E_{x}^{i}$, $I_{x}^{i}$, and $\tilde{I}_{x}^{i}$ considering endemic equilibrium. Administered severe patient data was not age-specified, but daily confirmed case data was. Therefore, we used confirmed case number by age data to estimate the initial states. Let [${DATA}_{i}$] is the age-specified data, then initial states are formulated as,

$$\sum_{k} I_{k}^{i}\left( 0 \right)=\frac{[DATA]}{\rho^{i}\alpha}$$

$$\sum_{k} E_{k}^{i}\left( 0 \right)=\frac{[DATA]}{\rho^{i}\kappa}$$

$$\sum_{k} \tilde{I}_{k}^{i}\left( 0 \right)=\frac{\left( 1-\rho^{i} \right)}{\rho^{i}}\frac{[DATA]}{\eta}.$$

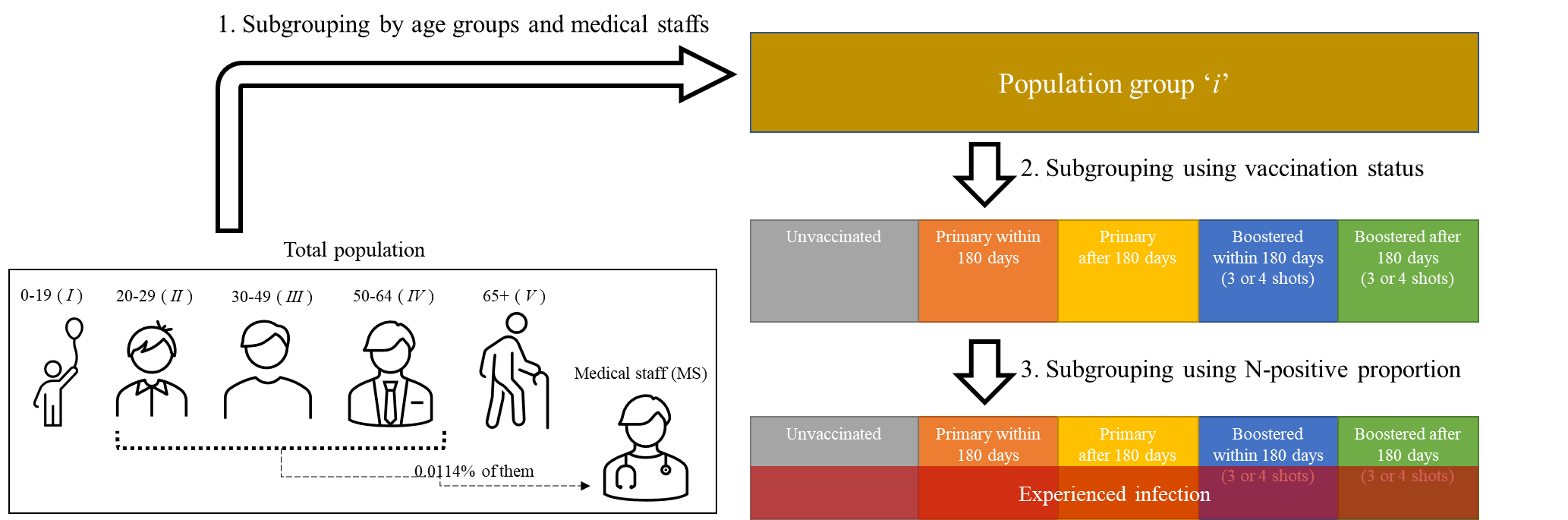

**Supplementary Figure** 1 Process of the population subgrouping for initial state of the model

**Supplementary Table** 3 Initial states of the susceptible groups

| **Prior infection** | **Age group** | **0-19** | **20-29** | **20-29 MS** | **30-49** | **30-49 MS** | **50-64** | **50-64 MS** | **65+** |
| --- | --- | --- | --- | --- | --- | --- | --- | --- | --- |
| No | Unvaccinated | 1,009,507 | 938 | 2,630 | 498,823 | 6,786 | 318,647 | 6,713 | 73,575 |
|  | Primary | 69,492 | 10,490 | 0 | 19,617 | 0 | 2,548 | 0 | 997 |
|  | Primary-waned | 629,970 | 724,788 | 0 | 1,173,948 | 0 | 362,240 | 0 | 124,754 |
|  | Booster | 372,171 | 739,628 | 11,835 | 1,626,625 | 30,535 | 2,468,874 | 30,207 | 3,540,766 |
|  | Booster-waned | 128,005 | 804,801 | 11,835 | 2,565,402 | 30,535 | 2,668,848 | 30,207 | 1,410,244 |
| Yes | Unvaccinated | 4,053,780 | 4,107 | 4,795 | 1,369,395 | 10,044 | 810,076 | 7,915 | 175,011 |
|  | Primary | 70,724 | 11,047 | 0 | 16,832 | 0 | 2,103 | 0 | 794 |
|  | Primary-waned | 1,502,253 | 1,831,454 | 0 | 2,133,284 | 0 | 620,958 | 0 | 203,150 |
|  | Booster | 175,340 | 357,198 | 21,578 | 674,763 | 45,197 | 994,157 | 35,618 | 1,387,988 |
|  | Booster-waned | 293,816 | 1,954,409 | 21,578 | 4,515,595 | 45,197 | 4,436,572 | 35,618 | 2,229,117 |

**Metropolis-Hasting algorithm results**

Supplementary Figure 2 shows the Markov Chain Monte-Carlo sampling results. Cumulative confirmed cases by age were used as fitting target data. We assumed Gaussian additive noise, i.e., variance was integrated out, and uniform prior distribution. Sample size was set as 40,000 per parameter, and we discarded first 7,000 samples from burn-in period. As results, samples of $h$, $\delta^{Ⅱ}$, $\delta^{Ⅲ}$, $\delta^{Ⅳ}$, $\delta^{Ⅴ}$, and $m_{2}$ had mode value as 0·5232, 1·7247, 0·9668, 1·5304, 2·7549, and 0·9307, respectively, and oscillated but had relatively narrow confidence interval, which is lesser than ${10}^{-4}$. Samples of $m_{3}$ ($m_{4}$) had mode value as 1·5086 (1·2650) and ranged from 1·4799 to 1·5297 (1·1705 to 1·3958) considering 95% of confidence interval. Mode values of the distributions were used for the model simulation.

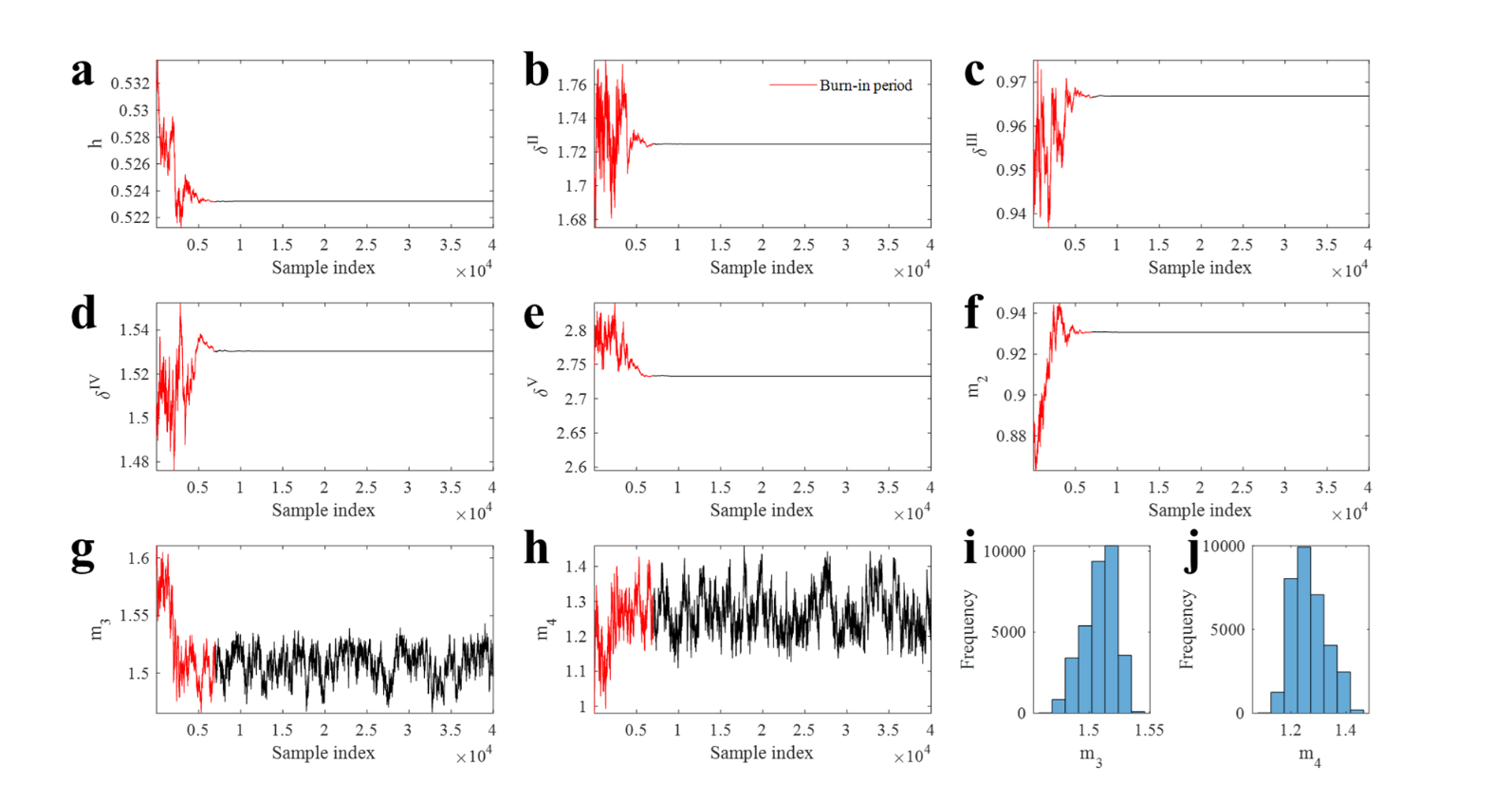

**Supplementary Figure** 2 Metropolis-Hasting algorithm application results. (a) to (h) Trace of the samples. (i) and (j) The posterior parameter distributions of the parameter $m_{3}$ and $m_{4}$, respectively.
